# The effect of alpha-cyclodextrin on postprandial glucose excursions, a systematic meta analysis *Alpha-cyclodextrin and Blood Glucose*

**DOI:** 10.1101/2022.09.04.22279468

**Authors:** Knut M. Wittkowski

**Author notes:** Correspondence: Knut M. Wittkowski, ASDERA LLC, New York, NY 10028, USA (KMW).

## Abstract

Alpha-cyclodextrin (αCD) is a natural bacterial product that is widely used as a food ingredient. In the EU, αCD is regulated as a “dietary fiber” with an authorized health claim “for contributing to the reduction of postprandial glycemic responses”. In the US, αCD is generally recognized as save (GRAS), but on April 25, 2022, the U.S. Food and Drug Administration (FDA) rejected the inclusion of αCD in the list of “dietary fibers” because “the strength of the scientific evidence does not support a finding of a beneficial effect of αCD on postprandial blood glucose …”.

This meta-analysis reviews clinical trials conducted to test the effect of αCD on blood glucose and insulin levels during three hours after consumption of a meal comprising carbohydrates, fats, and proteins.

Several issues related to the standardization of the outcomes, the choice of the statistical methods in the cross-over studies conducted, and the choice of methods for the aggregation of P-values are discussed. In conclusion, the EU decision is confirmed and extended. The administration of αCD not only reduces the postprandial glycemic responses, but the absence of an increase in insulin levels suggests that αCD acts independent of increasing insulin production and, thus, their beneficial effect is not affected by insulin resistance.

## INTRODUCTION

As soluble fibers, cyclodextrins (CDs) comprise several glucose molecules. In contrast to many other resistant fibers, however, CDs do form rings, rather than linear structures, which make them a prebiotic accessible only to some (commensal) bacteria, which have the enzymes to open these rings and digest the glucose units.

These rings have an interesting physicochemical property: they are hydrophilic on the outside, making them water-soluble, and lipophilic on the insight, allowing them to carry lipids through water (gut, serum), either to deliver lipid drugs or to filter out (“deplete”) lipids from gut or serum.

Alpha-, beta-, and gamma-CDs (αCDs, βCDs, γCDs) are rings of six, seven, and eight sugars, respectively, and, thus, can fit lipids of different size. αCDs bind preferentially saturated and trans fatty acids (FAs) [1-5], the larger βCDs can also fit steroids and sterols, while γCDs can carry even larger molecules. Their high specificity for “bad” FAs makes αCDs particularly interesting as dietary fibers.

In several clinical trials, αCDs have also shown to reduce the increase of blood sugar after a carbohydrate-rich meal. This meta-analysis aims to integrate the information from these clinical trials using standardized proportions of cyclodextrin vs carbohydrate or fat contained in the meals and standard statistical methodology for combining p-values in meta-analyses. Previous versions of this article were posted to the medRxiv preprint server on September 6, 2022 [6].

## METHODS

### Systematic review and meta analysis

#### Guidelines

This systematic review and meta-analysis is carried out in accordance with guidelines issued by Preferred Reporting for Systematic Reviews and Meta-analyses (PRISMA) [7].

#### Study selection

All articles identified from all the databases were imported into one Endnote library where all duplicates were removed. Studies that did not report the required data were excluded.

#### Data extraction

For each included article, the means, standard errors and p values related to incremental area under the curve (iAUC) were extracted as reported. Other extracted variables were age, gender, study duration, year of publication, and methodological characteristics.

### Statistics

When only *P*-values, but no effect estimates or individual data are available, meta-analyses can be conducted by aggregating information from *P*-values. Fisher’s combined probability test (FCPT) [8] is the most asymptotically optimal method in terms of Bahadur efficiency [9]. If results are concurrent (pointing into the same direction), the combined *P*-value can be calculated from:

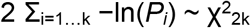

For instance, the requirement of two studies (*k* = 2) significant at the conventional *P*_*i*_ = 0.05 level for drug approval translates to a combined *P*(4×2.996 = 11.98 > χ^2^_4_) = .0175. One important consequence is that including an additional concurrent test with *P* < 0.37 (asymptotically) improves overall significance [10]. Hence, a third study with any *P* = 0.05 … 0.37 improves the overall significance of two studies conventionally significant at the 0.05 level each, rather than raising questions about their overall significance. If one of the two studies were significant at a level of e.g., P < .01, the third study might have to point into the opposite direction to void the significance of the other two studies.

When *P*-values are reported as categorial (*P* > 0.05), a meta-analysis can also be guided by the inspection of the figures in the source publications. To facilitate comparisons, the figures given below have been scaled and cropped, and elements have been resized or removed (like indicators of significance of comparisons not being considered), while some information have been added from the text. No data were removed, except values beyond 180 min (2 h) in two studies [4, 11].

## MATERIALS

### Literature Search

In September 2022, three publicly available data bases (clinicaltrials.gov, PubMed, and Google Scholar) were searched for clinical trials regarding the efficacy of αCD in reducing postprandial glucose excursions after a meal rich in carbohydrates. A manual search of the references lists of included articles and their citations was conducted to identify any articles not retrieved by the database searches.

### Inclusion and exclusion criteria

This systematic review and included clinical studies of dietary interventions that (1) enrolled, regardless of their age and background, who were generally healthy, (2) were randomized or non-randomized, (3) included carbohydrates in the meals, (4) included alpha-cyclodextrin as an intervention, and (5) included (postprandial) blood glucose profiles as an outcome. Many publications also included insulin and triglyceride (TG) profiles, but a search for “insulin” or “triglyceride” did not yield additional results.

### Records Retrieved

Data sources used where ClinicalTrials.gov, MedLine (PubMed), Google Scholar as well as the publications identified as relevant in these sources or referencing the sources (»).

<u>ClinicalTrials.gov</u> (Study type: Interventional Studies / Clinical Trials, Intervention/Treatment: Alpha-Cyclodextrin, Outcome Measure: Glucose, Other term: carbohydrate – removed to get results)

**Lytle, … Jensen (2018)** [11] NCT02999620 / NCT03002168

Amar *et al*. (2016) [12] NCT01131299 – 6 g/d, 12–14 wk, no single-day profiles taken

Soldavini *et al*. (2022) [13] NCT05393843 – terminated because of poor compliance because of coronavirus disease 2019 (COVID)

<u>MedLine</u> (Clinical Trial, “carbohydrate alpha-cyclodextrin glucose”)

**Buckley *et al*. (2006)** [14]Comerford *et al*. (2011) [15] – glucose profiles not an outcome

» Grunberger *et al*. (2007) [16] – serum glucose measured in diabetic patients not reported

**Gentilcore *et al*. (2011)** [4]

**Jarosz, Flexner *et al*. (2013)** [17]

Amar *et al*. (2016) – <u>duplicate</u>

Bessell et al. (2020) [3] – glucose profiles not an outcome, deviation from study protocol (“dietary advice for weight loss”) [18]

<u>Google scholar</u> (carbohydrate alpha-cyclodextrin glucose postprandial clinical)

Buckley *et al*. (2006) – <u>duplicate</u>

Gentilcore *et al*. (2011) – <u>duplicate</u>

Jarosz, Fletcher, *et al*. (2013) – <u>duplicate</u>

Amar (2016) *et al*. – <u>duplicate</u>

Jain *et al*. (2016) – glucose profiles are not an outcome.

**Bär, Diamantis, *et al*. (2020)** [5] – in PubMed, but not as a “clinical trial”, unpublished protocol (2002) and report (2002) quoted in EFSA (2012) [19] some results also described in **Schmid (2004)** US 2004/0161526 A1 (Wacker, abandoned) [20]

**» Sugahara, Inoue, *et al*. (2016)** [21] – not in PubMed, not on Internat J Pharmacy Web site

Lytle, …, Jensen (2018) – <u>duplicate</u>

**Binou *et al*. (2022)** – not in PubMed

The PRISMA 2020 Flow Diagram [7] is shown in Figure 1.

**Figure 1:**
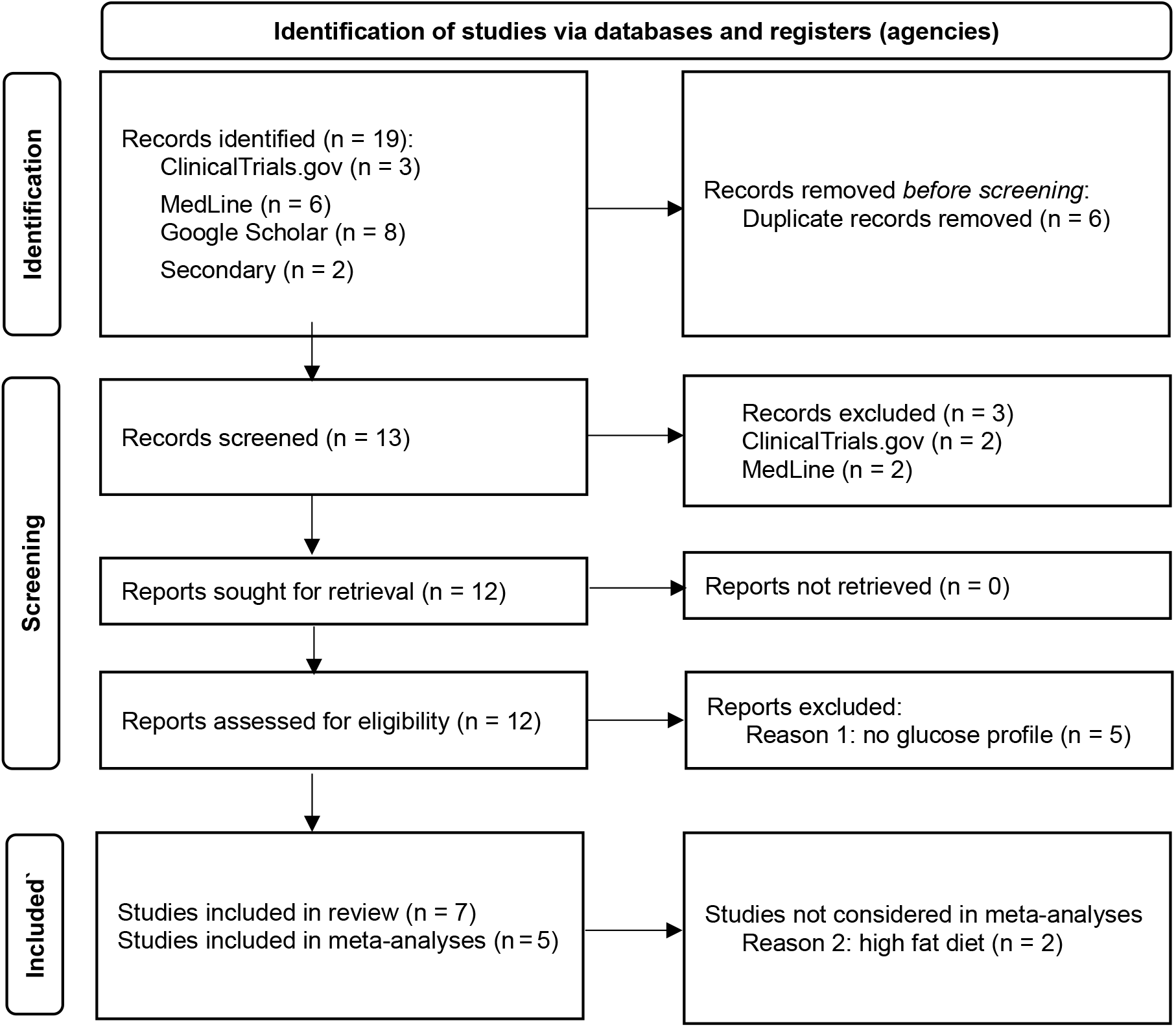
PRISM Flow Diagram: Preferred Reporting Items for Systematic Reviews and Meta-Analyses (PRISMA) 2020 **[7]**

Three additional publications were identified. One had glucose profiles only as a secondary outcome [11], the other two were published only after the FDA had made its decision; one is included [22, 23], another study (a combination treatment with 5 g αCD per meal, 45% calories from carbohydrates) was terminated because of poor compliance during COVID [13].

The EFSA based it’s 2012 scientific opinion on the following three clinical trials:

**Buckley** *et al*. (2006) [14]:10 healthy adults age 24 ± 4 yr consumed boiled rice (50 g digestible carbohydrates) with 0, 2, 5, or 10 g of added αCD. Postprandial glucose and insulin were assessed.

**Diamantis, Bär** (unpublished 2002 protocol and report, quoted in EFSA (2012) [19], subsequently published as Bär, Diamantis, *et al*. (2020) [5]): 12 healthy male adults age 23–24 consumed white bread (50 g starch) with 0 or 10 g αCD dissolved in 250 mL drinking water. Postprandial glucose and insulin were measured. The study was also included in the patent application Schmid (2004) [20]

**Gentilcore** *et al*. (2011): 10 healthy older subjects age 68–78 yr consumed 100 g sucrose with 0 or 10 g aCD dissolved in water. Postprandial glucose and insulin were measured.

The FDA included two more recent studies, where the meals, in particular in the former study, also contained substantial amounts of fat and protein:

**Jarosz, Fletcher**, *et al*. (2013) [24]: a dissertation (Fletcher 2013) [24] published subsequently as Jarosz *et al*. (2013) [17]: 34 healthy adults age 18–65 yr consumed a commercially prepared egg sausage biscuit sandwich (32 g carbohydrates, 26 g fat, 20 g protein) with 0 or 2 g of αCD (2 pills). Postprandial glucose and triglycerides were measured.

**Sugahara, Inoue**, *et al*. (2016) [21]: 10 adults age 22.9±1.8 consumed a beef curry and rice meal (86 g carbohydrate, 13.5 g fat, 11 g protein) with 0 or 5 g αCD. Postprandial glucose, triglycerides, and insulin were measured.

This meta-analysis also contains the results from two studies reviewed by neither of the agencies:

(6) **Lytle, …, Jensen**, *et al*. (2018) [11]: 8 healthy adults age 23–54 yr consumed 2 g αCD with a liquid meal breakfast comprising 60% (37–54 g) carbs, 27.5% fat, and 14.5% protein. While the primary outcome was fat, plasma glucose was also measured.

(7) **Binou** *et al*. (2022) [22, 23] ^NCT04725955^: 10 healthy adults age 18–41 (28.2±6.8) consumed white wheat bread (50 g carbohydrates, 2-4 g fat, 10–12 g protein) with 0 or 5 g of αCD. Postprandial glucose and insulin were measured.

## RESULTS

### Data Extraction

Table 1 shows the numerical results extracted from the seven studies included in the review.

**Table 1:** Summary Data.

| Agency | Publications | Sub-jects | Age [yr] | Ingredients | Source | αCD [g],<br>(... / 50 g) | glucose | insulin | TG |
| --- | --- | --- | --- | --- | --- | --- | --- | --- | --- |
| EFSA, | Buckley et al. (2006) [14] | 10 | $24 \pm 4$ | 50 g starch | white rice | 2, (2) | ↓ n.s. | ↑ n.s. | |
| FDA | ... | healthy | ... | ... | ... | 5, (5) | ↓ .03 | ↔ |  |
| ... | ... | adults | ... | ... | ... | 10 (10) | ↓ .001 | ↓ n.s. |  |
| EFSA,<br>FDA | Diamantis (2002a/b referred to in in EFSA 2012) [19], Bär (2020) [5], Schmid (2004) [20] | 12 healthy male adults | 23–24 | 50 g starch / ~1 mL olive oil | White bread | 5 (10) | ↓ <.05 | ↓ <.05 |  |
|  | Binou et al. (2022) [22, 23] | 10 healthy adults | 18–41 | 50 g carbs / ~3 g fat / ~11 g protein | white wheat bread | 5 (5) | ↓ <.06 | ↔ |  |
| EFSA,<br>FDA | Gentilcore et al. (2011) [4] | 10 healthy seniors | 68–78 | **100 g sucrose** |  | 10 (5) | ↓ n.s. (1h: <.05) | ↓ .09 (2h: <.001) |  |
| FDA | Sugahara et al. (2016) [21] | 10 healthy adults | $23 \pm 2$ | **86 g carbs / 13.5 g fat / 11 g protein** | beef curry with rice | 5 (3) | ↓ <0.37 | ↓ n.s. | ↓ n.s. |
| FDA | Fletcher (2013), Jarosz et al. (2013) [17, 24] | 34 healthy adults | 18–65 | **32 g carbs / 26 g fat / 20 g protein** | McMuffin egg sausage biscuit | 2 (3) | ↓ n.s. |  | ↓ <.05 |
|  | Lytle, ..., Jensen et al. (2018) [11] | 8 healthy adults | 23–54 | ~45 g carbs, **~21 g fat** ~11 g protein | Ensure Plus liquid meal | 2 (2) | ↔ |  | ↔ |

Figure 2 shows the average glucose profile data by diet as presented in the seven clinical trials. Figure 3 shows the serum insulin or triglyceride (TG) profiles as provided in the publications.

**Figure 2:**
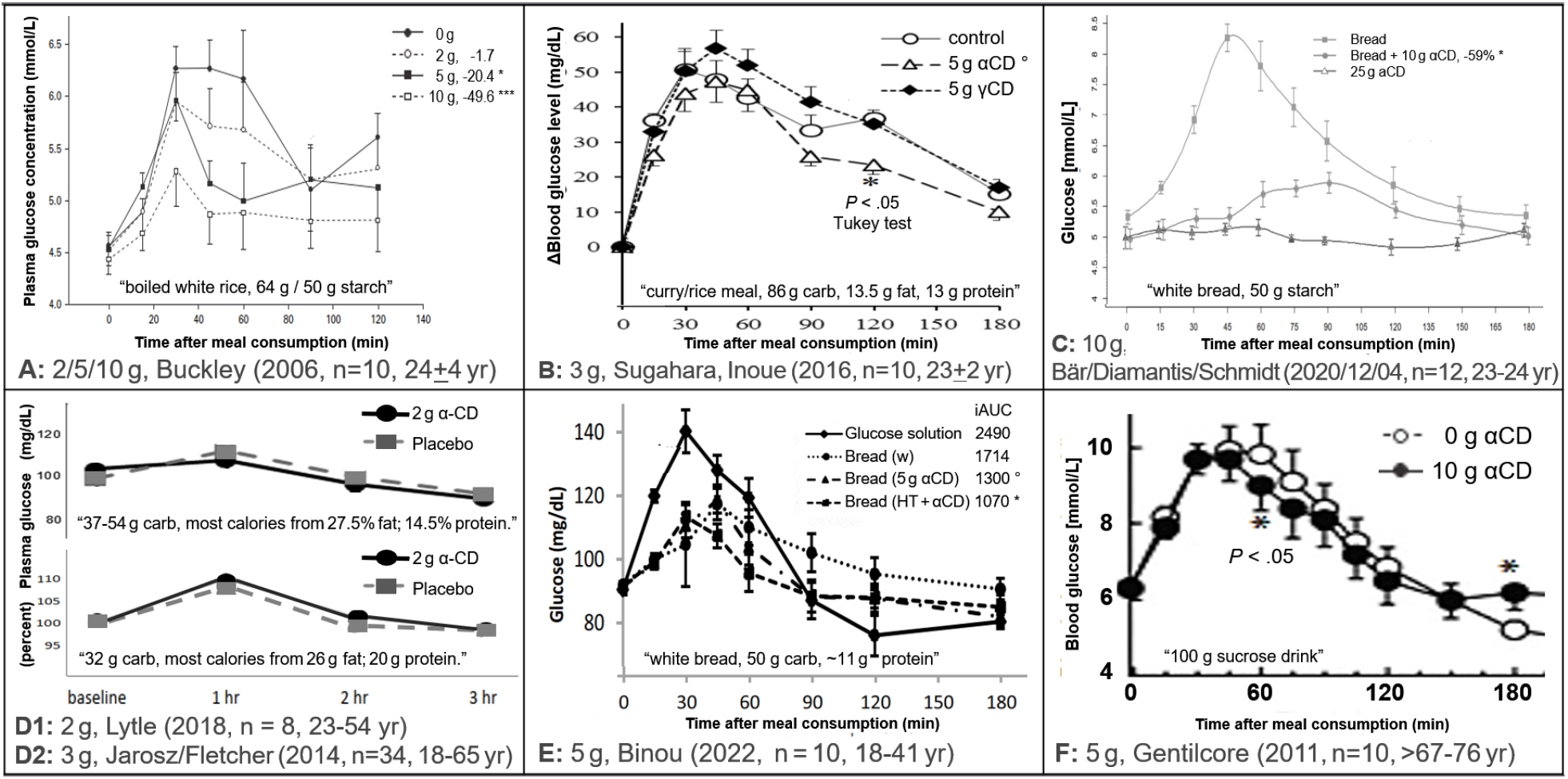
Data Extraction Summary: Effects of different doses of αCD on blood glucose profiles. In all studies, each of >8 patients were administered all doses (cross-over). ***: *P* < .001; *: *P* < .05; °: *P* < .37, Figures were derived (excerpted) under 17 U.S.C. § 107 (fair use in scholarship and research) from Buckley *et al*. (2006, permission granted) [14], Sugahara, Inoue, *et al*. (2016) [21], Bär, Diamantis, *et al*. (2020, CC BY 4.0) [5] Fletcher, *et al*. (2014, dissertation) [17], Lytle *et al*. (2018, redrawn) [11], Binou, *et al*. (2022, permission granted) [22, 23], Gentilcore, et al. (2011, permission granted) [4]: To serve the purpose of scholarly criticism, Figures have been scaled to have similar axes, scales were cropped at 180 min [4, 11], colors and some misleading indicators of “significance” were removed, and information was added to their sub-legends.

**Figure 3:**
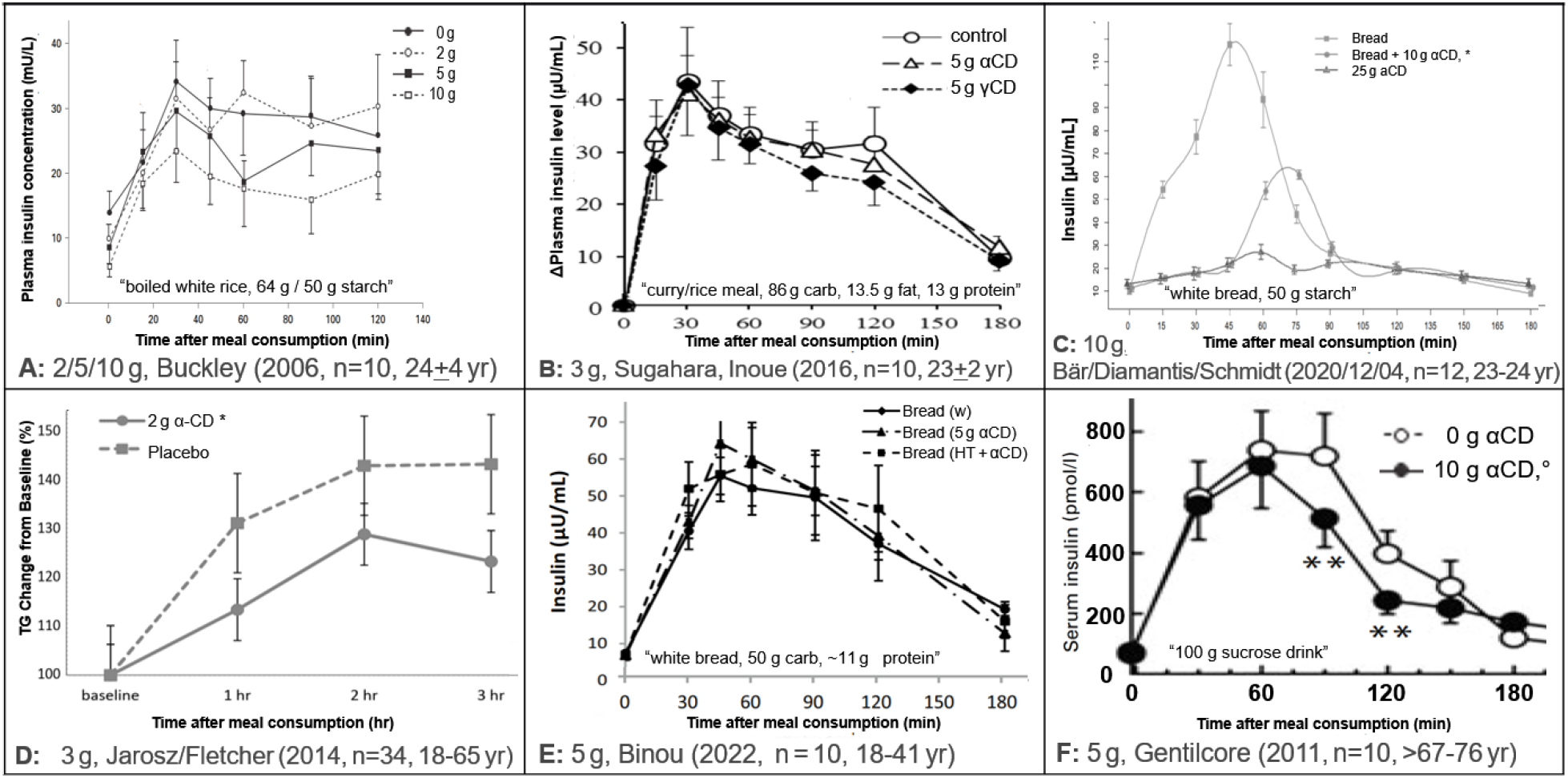
Data Extraction Summary: Effects of different doses of αCD on serum insulin and triglycerides (TG). See Figure 2 for legend.

### Data Review

One of the most striking observations are the glucose curves in the two studies with the highest fat and protein content [11, 17], where blood glucose levels increased by only 10% under control conditions, compared to ∼50% in studies serving a high-carbohydrate diet. (Figure 1D has been rescaled from [24, Figure 2] to reflect that aspect.) If a diet does not increase blood glucose, there is no increase that αCD can significantly reduce, especially not at a small sample size. Hence, these studies will not be considered for the meta-analysis.

Since the earliest study [14], the use of standard error of mean (SEM) bars shown in the graphs point to another, potentially common problem in this field: the data collected under different conditions seem to have been analyzed by statistical methods that ignore the within-subject correlations (WSC), which is a key element in the analysis of a cross-over design. Ignoring the WSC seems to have contributed to several of the results being reported as “non-significant” despite obvious differences seen in the figures. The text in another study [21], for instance, reads “the cumulative [incremental area under the curve] iAUC … was smaller in the αCD group … than in the control group”. This strongly suggests that the results are described as if the data were coming from different “groups”, so that the WSC was ignored, diminishing the reported level of significance. In the latter study, their Fig. 3 caption (here, Figure 2B) notes that “Tukey’s test” was used. Assuming that this refers to Tukey’s (1949) test for “comparing individual means in the analysis of variance” [25], this test is not appropriate for comparing the 0–180 min cumulative iAUC.

### Meta Analysis

Among the studies with high levels of carbohydrates (bread and rice), the earliest study [14] (Figure 1A) noted “a dose-dependent inhibition of the postprandial glycaemic response to a standard carbohydrate meal following incorporation of [αCD]. The mean iAUC for glucose was negatively related to the dose of α-CD (*r*^2^ = 0.97, *P* = 0.02), with the iAUC being significantly lower than the control dose (0 g αCD) for the 5-(*P* = 0.03) and 10-gram (*P* = 0.001) doses”.

The comparisons based on the four subsequent studies [4, 5, 21-23], excluding the two studies serving high-fat meals [11, 17], are consistent with a dose-response relationship [14]:

**2 g / 50 g:** As in the initial study [14], the 2 g dose was not significant: **No evidence for efficacy**.

**3 g / 50 g:** One study [21] served beef curry containing fat with 86 g carbs per meal, so the effective “5 g” dose was only 2.9 g per 50 g of starch. Moreover, the Fig. 3 bars being obviously, but not “significantly” different strongly suggests at least one of the above statistical flaws.

**Glucose: efficacy is unclear because of statistical issues**; the publication is not included on the Internat J Pharm Web site [21].

**Triglycerides:** In a second study [17], where subjects were fed a high-fat (26 g) McMuffin egg sausage biscuit, 2 g αCD had **significant efficacy** (*P* < 0.05).

**5 g / 50 g:** In the initial [14] and a second study [22, 23], 5 g were significant (at *P* = 0.03 and

*P* = 0.06, respectively, combined ***P* = 0.013** [8], well below the FCPT cut-off of 0.0175).

One of the “10 g” studies (actually: 5 g per 50 g sucrose) is excluded from this meta-analysis because the participants were older (68–76 yr) and consumed a sucrose drink [4], rather than a high-carb meal. Still, at least one significant difference between “groups” suggests a *P* < .37, so that including the results should increase the significance from the other two studies.

**Glucose: there was significant efficacy from at least two studies** [14, 22, 23]. **Insulin:** The reduction in one study [4] was not significant (*P* = 0.09).

**10 g / 50 g**: The 10 g dose was significant in the initial [14] and in a second study [5] (*P* < 0.001 and *P* < 0.05, respectively, combined ***P* < 0.00055**) [8].

**Glucose: there was highly significant efficacy from two studies** [5, 14].

**Insulin:** efficacy was significant (*P* < 0.05) in the one study with data [5].

In summary, all comparisons, even at the 2 g dose, showed lower glucose profiles with αCD and only one comparison (at the 2 g dose) [4] showed higher (but not significant) insulin levels with αCD. Hence, the efficacy of αCD when added to a high-carb (rice or bread) meal in young adults is dose-dependent [19], with doses from **5 g/50 g** proven effective, as concluded by the EFSA in 2012. The αCD’s mechanism of action seems not to involve an increase of insulin production.

## DISCUSSION

This meta-analysis addresses one of the hypothetical benefits of αCD: the reduction of postprandial glycemic responses, where EFSA [19] and FDA [26] have come to different conclusions, even though they agree on the interpretation of the three studies available to the EFSA in 2012: (1) 50 g starch in white rice, 2/5/10 g αCD, young adults αCD, young adults [14]: “<u>a dose-dependent effect</u>” [19], with no effect at 2 g, but “significant effects of <u>5 g and 10 g</u>” [26].(2) 50 g starch in white bread, 10 g αCD, young adults [5]: <u>a (significant) effect</u> [19, 26]. (3) 100 g sucrose drink, 10 g αCD, seniors [4]: no effect [19, 26].

### EFSA

“The Panel considers that the following wording reflects the scientific evidence: “Consumption of alpha-cyclodextrin contributes to the reduction of the blood glucose rise after starch-containing meals” and “in order to obtain the claimed effect, at least 5 g of alpha-cyclodextrin per 50 g of starch should be consumed” [19].

Two additional studies had been published at the time of the FDA’s review (see Materials) [26]. (1) In a high-fat meal with 32 g starch, 2 g αCD taken by adults [17] didn’t show any significant difference [26]. This study was excluded from the meta-analysis, because of the diet’s low carbohydrate content. After this high fat meal, however, αCD reduced TG. (2) In a beef curry/rice meal containing 86 g starch, 5 g αCD taken by adults [21] (2.9 g per 50 g of starch) didn’t show any statistical difference, either [26]. Aside from a low dose of αCD, the latter study raises substantial issues regarding the validity of the statistical analysis, as discussed above.

### FDA

Based on these studies, the FDA concluded that “there is inconsistent evidence … which weakens our confidence …” [26].

Another informative study has been published since: 50 g starch in white bread, 5 g αCD, *P* = 0.06 (< 0.37) [22, 23]. Despite being “not significant” on its own, this study clearly adds further evidence for the dose of 5 g αCD, when added to a high carbohydrate meal, being effective.

From the Results, there are several aspects in the published evidence that allow one to resolve at least some of the perceived inconsistencies.

### Lack of significance

In a meta-analysis, a “non-significant” result should not be interpreted as evidence against an effect. (“Lack of proof of an effect is not proof of lack of an effect”.) In particular, having one of several studies not reaching the conventional level of significance (*P* < 0.05) does not, in itself, create an inconsistency. With the FCPT [8], adding a study with P < 0.37 [21-23] typically suffices to strengthen the overall significance [10].

### Within-subject correlation

Cross-over studies often have more power than studies comparing groups because data coming from the same person tend to be correlated. At least one of the studies [21], however, apparently analyzed the data without accounting for that correlation, which may have contributed to the “non-significant” statistical results despite the published Figures showing clear evidence for an effect. Luckily, the *P*-values, in that study, were still below the *P* < 0.37 limit and, thus, contribute to the overall significance when summarized by the FCPT.

### High-fat vs high carbohydrate

One of the recent studies [17] with inconsistent results assessed the effect of αCD when added to a high-fat (rather than high-carb) meal, and, consequently, blood glucose levels did not increase to levels comparable to high-carb meals. Instead, triglyceride (TG) levels increased, and αCD exerted its beneficial effect by reducing the increase in serum TGs.

As another important issue, αCD may have other benefits with high-fat meals than reducing postprandial glucose excursions. In the study with the low-carb/high-fat meal [17], excluded from this meta-analysis for that reason, “consumption of α-CD with a fat-containing meal was associated with a significant reduction in postprandial TG responses” [17]. This significant result was supported by the potentially “nonsignificant trend” in the study with a fat-containing meal [21].

Hence, the overall benefit of αCD is not restricted to reducing the postprandial glucose response seen after carb-rich meals. With fat-rich meals [17], it is blood lipids (incl. TG) that increase and αCD reduces this response, instead of the glucose response after a carbohydrate-rich meal. In fact, much of the effect of αCD helping obese people to lose weight may be related to αCD reducing lipid, rather than glucose response. The benefit of αCD helping with controlling body weight has been demonstrated in several clinical trials: (1) Grunberger *et al*. (2007) [16] and Jen (2013) [1] showed that in obese people with type-2 diabetes age >30 yr and 57.5±9 yr, respectively, “αCD reduced body weight,” although this was “only significant after adjusting for energy intake”. (2) Comerford *et al*. (2011) [15], showed that in overweight people with type-2 diabetes age 41±13.6 yr “αCD significantly decreases body weight and [low-density lipoprotein] LDL” (3) Amar *et al*. (2016) [12] showed that in healthy subjects age 34±12.4 yr, αCD reduced small-LDL by 10% (*P* < 0.045) and, consistent with the results of this meta-analysis, reduced insulin resistance by 11% (*P* < 0.04)

Finally, the FDA noted that there was no clear relationship between postprandial insulin and postprandial glucose. A lack of such relationship, however, can, in fact, be an advantage, because it shows that αCD does not lower glucose by increasing insulin production and, thus, have the desirable effect on blood glucose even in people whose insulin production is impaired.

Weaknesses of this meta-analysis include the lack of subject-level data and often even of exact *P*-values. Another shortcoming is that the majority of studies were conducted in young adults, while seniors could potentially benefit the most. More studies in older populations are urgently needed.

## CONCLUSIONS

This meta-analysis is based on five published clinical trials on the efficacy of αCD to reduce glucose excursions after a high-carb meal containing ∼50 g starch from white bread or rice in subjects aged 18–41 yr. This effect does not seem to require an increase in insulin production. Two additional publications had the majority of calories came from fat. Hence, these two studies were excluded from the meta-analysis.

There were strong indications that the published *P*-values suffered from loss of power due to errors in the statistical methodology, including the cross-over design not being reflected in the statistical method used. Still, a formal meta-analysis using the FCPT confirmed the European Medical Agencies (EMA)’s 2012 health claim that, when taken with a high-carb meal, at least 5 g of the dietary fiber αCD per 50 g of carbohydrates reduce postprandial glucose excursions by a mechanism-of-action unaffected by insulin resistance.

## Data Availability

All data used are available online as part of the publications cited.

## Notes

### Competing Interest Statement

Author Knut M. Wittkowski is currently the CEO of ASDERA LLC, a company discovering novel pharmacological and nutritional interventions against complex (incl. cardiovascular) diseases from data of genome-wide association studies. The discussion of statistical aspects in the meta-analysis / systematic review in this publication was not affected by any commercial or financial relationships that could be construed as a potential conflict of interest.

### Funding Statement

This study did not receive any funding.

### Summary of Updates

copyright information added and text revised.

